# Evaluation of the Weightage on Pain Management in Pharmacology Curricula of the Undergraduate Medical Program (MBBS) of Bangladesh

**DOI:** 10.1101/2021.05.05.21256500

**Authors:** Jannatul Ferdoush, Fatema Johora, Fatiha Tasmin Jeenia, Tashfia Momtaz, Sharif Mohammad Towfiq Hossain, Halima Sadia, Asma Akter Abbasy, Md. Sayedur Rahman

## Abstract

**Background:** Prioritizing problem-oriented undergraduate medical education is paramount to adequate management of pain in real life scenarios. The present research was conducted with an attempt to explore the important baseline information for pain medicine education and evaluation within undergraduate pharmacology curricula in Bangladesh.

**Materials and methods:** This descriptive cross-sectional study evaluates the curriculum (pharmacology portion of undergraduate medical curriculum), and written question (SAQ) of MBBS Examination of last ten years extending from January 2010 to November,2019 of all 7 universities offering MBBS degree. The evaluation was conducted through searching certain key phrases.

**Result:** In Pharmacology & Therapeutics portion of the curriculum, only 4 hours and 2 hours are allocated to discuss pain management in lecture and tutorial respectively. In the study period, average marks allocated in pharmacology written question papers was 4.4 (*SD* = 2.7) and the difference among studied universities was not significant (p value 0.7).

**Conclusion:** Allocated time in the curriculato teach pain management is very low and weightage received in assessment is also inadequate. Education on pain medication as well as management should receive more emphasis.

## Introduction

Millions of people around the world suffer from acute and chronic pain due to inappropriate pain management that leads to significant dysfunction and disability, as well as a rising health care burden [1].In public hospitalsof Bangladesh, it is reported that over 400,000 patients admitted for road traffic accidents and assaults that topped the list of reasons for admission [1]. Acute pain is one of the most common reasons why people seek to the emergency department, and it’s still a concern in the post-operative setting [3].If an acute pain dealt inadequately, it transforms into chronic pain making the individual a burden to the society [4].It is critical for future medical graduates to be competent in the field of pain medicine in order to prepare them for the real-life situation [5].

One of the barriers to best-practice pain management has been identified as a lack of knowledge and training in the field of pain medicine [6].According to a survey conducted in developing countries by International Association for the Study of Pain (IASP) revealed substantial shortcomingson pain education and clinical training.Over 90% medical graduates reported that the education on pain was insufficient to meet the necessity of their practice [7,8].Most medical school curricula allocate little time to pain education and that is also not incorporated into case-based clinical practices [7].

Allocating additional time for pain to an already compact medical curriculum mean compromising with some other content [5].This makes it difficult for introduction of new content into the undergraduate medical curricula. However, it has been more than 30 years since the International Association for the Study of Pain (IASP) first published a comprehensive “core professional curricula” for teaching on all aspects of pain[9].Despite its publication over 30 years ago, significant gaps persistat the undergraduate level [8].Consequently,postgraduate trainees and practicing physicians struggle with competency[5].There is also a lack of integration of basic science and clinical expertise. Although students might be academically sound, they remain unsuccessful in the clinical application of that knowledge, perhaps, due to a dearth of training [4].

Now-a-days, most medical school curricula around the world have a dearth of pain-related content[10-12].The treatment of pain necessitates a multidisciplinary approach, but pain management is taught to undergraduates in limited clinical classes[4].

More than a decade ago, the Joint Commission on Accreditation of Healthcare Organizations published guidelines to ensure that healthcare professionals are properly qualified to treat acute pain [13]. Current methods of delivering pain education should focus on an integrated approach for students to make them capable to apply their taught lessons effectively and confidently in treating pain as professionals[10].

To reduce the burden of pain in society, undergraduate curricula should include sufficient training, education, time, and research [14, 15].On this background, the present study has attempted to analyze the academic documents (curricula, and question papers) regarding pain education. The aim of the study was to evaluate the status of pain management education in Bangladesh. This study is expected to enable the researcher to get a precise concept about the current situation regarding pain management and analgesia issue in undergraduate medical education of Bangladesh.

## Materials and Methods

### Study Design and Procedure

The study was a descriptive cross-sectional one and was conducted from January, 2021 to March, 2021. Undergraduate curricula (pharmacology portion) and written question papers of 7 (seven) universities were reviewed with method adopted from a previous study [16].

### Selection of key phrases relevant to postoperative pain management

In order to review the curricula and written question, the principle of conceptual mapping was applied by a panel of experts (3 senior Pharmacologists) and the following key phrases were identified: pain, analgesia, analgesic, analgesic property, postoperative pain, pain management, visual Analog Scale (VAS), analgesic ladder, analgesic guideline, opioids, narcotic, non-narcotic, non-opioids, NSAIDs, COX I, COX II, morphine, pethidine, tramadol, codeine, aspirin, diclofenac, paracetamol, ketorolac, ketoprofen, indomethacin, naproxen, cewlecoxib, rofecoxib, etorocoxib.

#### Undergraduate Medical Curricula (MBBS)

Undergraduate medical curricula was searched for the mentioned key phrases in the soft copies of the curricula of MBBS (Pharmacology& Therapeutics portion), and then the area was identified, where the key phrases were mentioned.

#### Written Question Papers of MBBS Examinations

Pharmacology written question papers (SAQ) of MBBS Examination of last ten years (Jan 2010 to November-2019) of all 7 universities offering MBBS degree (University of Dhaka, University of Chittagong, University of Rajshahi, Shahjalal University of Science and Technology, University of Science and Technology, Chittagong, Bangladesh University of Professionals and GonoBishwabidyalay) were collected and analyzed. Same key phrases were searched and the number of occasions they appeared in question paper was calculated and weightage given in written questions were assessed.

## Result

**Table I** shows that out of total 200 hours of teaching in Pharmacology & Therapeutics portion of the MBBS curricula, 4 hours and 2 hours are allocated to discuss pain management in lecture and tutorial, respectively.

**Table I:** Allocated Teaching Hours for the Topics Those Mentioned Selected Key Phrases in Undergraduate Medical (MBBS) Curricula of Pharmacology.

| Key Phrase | Teaching Learning Strategies (allocation in hours) |  |  |  |  |
| --- | --- | --- | --- | --- | --- |
|  | Lecture | Tutorial | Practical | Clinical Case Reports | Total |
| Total allocation (in hours) | 100 | 30 | 50 | 20 | 200 |
| Allocation on Pain | 04 | 02 | 0 | 0 | 06 |
| Opioid analgesics | 02 | 0 | 0 | 0 | 02 |
| NSAIDS/ Non-opioid analgesics | 02 | 02 | 0 | 0 | 04 |

**Table II** shows the mean allocated marks in the pharmacology written questionsin undergraduate medial program of different universities during the study period extending from January 2010 to November 2019. The average allocated mark was 4.4±2.7 and the difference between the universities was not statistically significant (p value 0.7).

**Table II:**
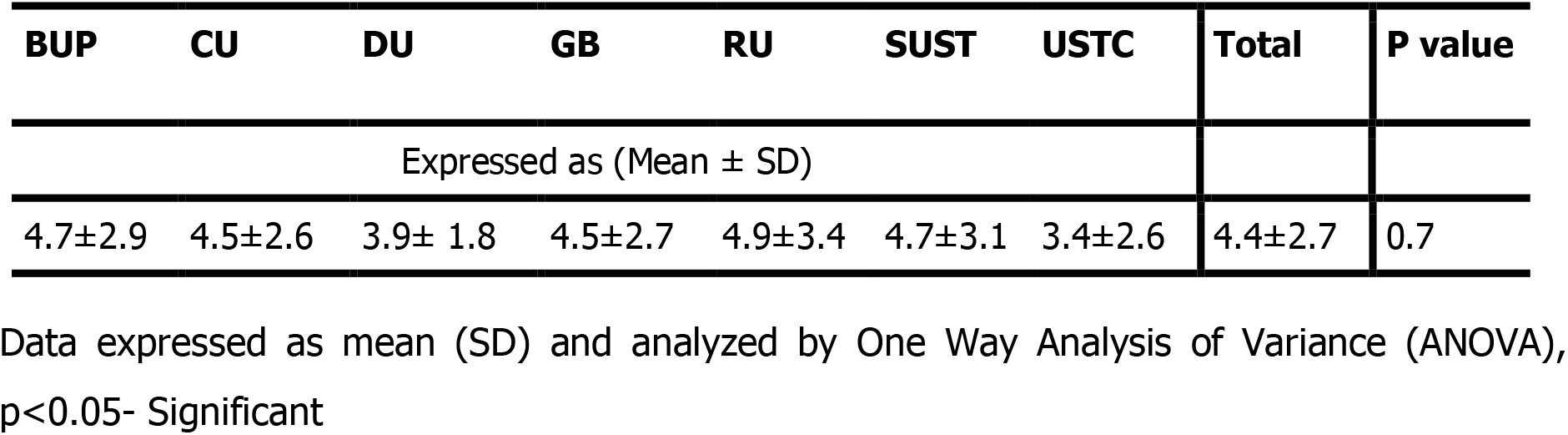
Written Questions of Pharmacology in Undergraduate Medical Program (from January 2010 to November 2019)

| BUP | CU | DU | GB | RU | SUST | USTC | Total | P value |
| --- | --- | --- | --- | --- | --- | --- | --- | --- |
| Expressed as (Mean ± SD) | | | | | | | | |
| 4.7±2.9 | 4.5±2.6 | 3.9± 1.8 | 4.5±2.7 | 4.9±3.4 | 4.7±3.1 | 3.4±2.6 | 4.4±2.7 | 0.7 |

Data expressed as mean (SD) and analyzed by One Way Analysis of Variance (ANOVA), $p < 0.05$ - Significant

BUP: Bangladesh University of Professionals; CU: University of Chittagong, DU: University of Dhaka, GB:GonoBishwabidyalay RU: University of Rajshahi, SUST: Shahjalal University of Science and Technology, USTC: University of Science and Technology (USTC),

**Figure I** showed trend of weightage on pain medicine in DU and CU in study period. In DU, there was gradual increase of weightage but sharply declined from 2015-17, later a spike was observed in 2018 and again declined in 2019. On the other hand, in case of CU, constant weightage was observed in 2010-11 but there was zero weightage in 2012, then gradual increase of weightage was found from 2013-16, a spike was observed in 2018 and again declined in 2019.

**Figure 1:**
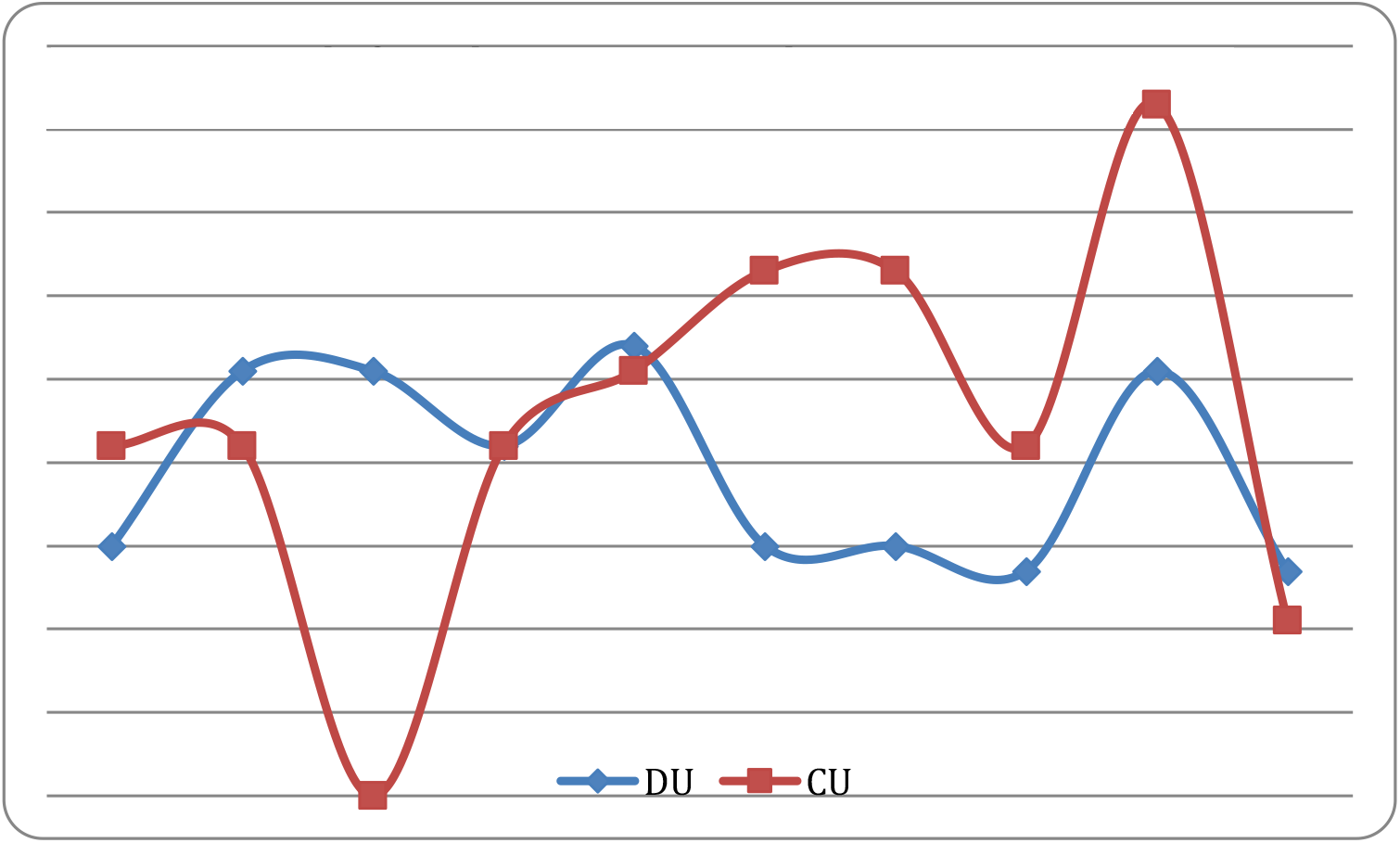
Trend of weightage on pain medicine in University of Dhaka (DU) and University of Chittagong (CU) during study period.

**Table III** shows content of pharmacology curricula covering pain medications in undergraduate medical curricula. Mainly drugs name, classification and mechanism of action, clinical use and adverse effects mentioned in the content.

**Table III:**
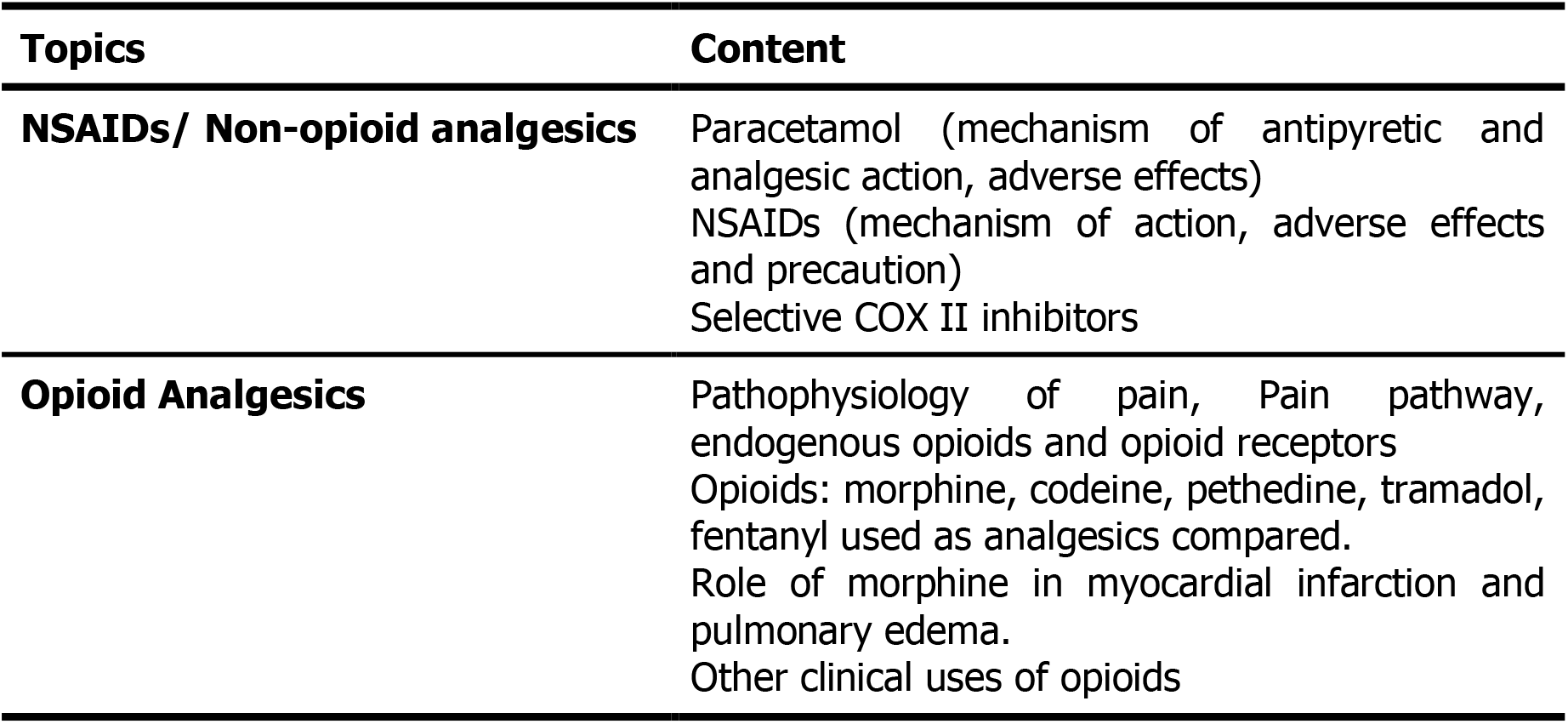
Content Covering Pain in the Pharmacology portion of Undergraduate Medical Curricula.

| Topics | Content |
| --- | --- |
| **NSAIDs/ Non-opioid analgesics** | Paracetamol (mechanism of antipyretic and analgesic action, adverse effects)<br>NSAIDs (mechanism of action, adverse effects and precaution)<br>Selective COX II inhibitors |
| **Opioid Analgesics** | Pathophysiology of pain, Pain pathway, endogenous opioids and opioid receptors<br>Opioids: morphine, codeine, pethedine, tramadol, fentanyl used as analgesics compared.<br>Role of morphine in myocardial infarction and pulmonary edema.<br>Other clinical uses of opioids |

## Discussion

Pain education now constitutesa minor topic in undergraduate medical programs, as evidenced by the limited time allotted to pain-related topics. This study attempted to assess the weightage on pain medicine in undergraduate pharmacology education of Bangladesh.

The present study revealed that the undergraduate Pharmacology & Therapeutics curriculum only devotes 4 hours in lecture and 2 hours in tutorial to teach pain management.Studies conducted in many developed countries, such as the United States [12], Canada [17],Norway [18],and the United Kingdom [10] demonstrates higher average hours dedicated to pain education.Shipton et al. (2018) found that time allotted for teaching pain medicine ranged from 5 to 43 hours, with an average of 19.6 hours during entire medical curricula.[22].These findings pointed out that pain therapy receives insufficient focus in medical education disproportionate to the clinical and social burden of pain [20].Moreover, according to a recent study, veterinary scientists place a higher value on undergraduate pain education than do healthcare professionals [14].

Through examination, whether the objectives are met or not is evaluated[21]. Written exams were used to assess student knowledge in pain medicine. These approaches are not appropriate to assess skill-based pain competencies and attitudes.

The current research revealed that in pharmacology written question papers average allocated marks in the study period was 4.4±2.7 indicating that pain medications have received insufficient attention and therefore received low priority. Shipton et al. (2018)also identified that assessment methods are predominantly MCQs and short-answer questions. [22].

The present study found that Opioid analgesics and NSAIDS are the main pain related topics in pharmacology curricula with content focusing on classification, drug names, and mechanism of action, clinical use, and adverse effects. Whereas, psychological pain management, ethics and medicolegal dimensions, multidisciplinary pain clinic, geriatric painand pediatric pain are overlooked[12].

The amount of time and resources allotted to a subject, as well as its weightage during assessments, are principal determinant of its importance. This study also revealed the trend of weightage on pain education in two leading universities of Bangladesh, which steadily increase in 2018 but then decline in 2019. According to this finding, the lack of attention paid to assessing pain medicine competencies suggest a general lack of importance placed on the subject by medical universities.

## Conclusion

The emphasis on pain education in the curricula of undergraduate medical education is out of proportion, given the burden of pain in the general population in Bangladesh.More time should be allocated for teaching painand methodologies should be modified to facilitate problem-solving, deeper learning and skill development. The results of this study will serve as the ignition at the conceptual level of pain management education, as well as expected to facilitate institutions to review their content and redesign their curricula.

## Data Availability

This descriptive cross-sectional study evaluates the curriculum (pharmacology portion of undergraduate medical curriculum), and written question (SAQ) of MBBS Examination of last ten years extending from January 2010 to November,2019 of all 7 universities offering MBBS degree. The evaluation was conducted through searching certain key phrases.

## Reference

1. Stewart WF, Ricci JA, Chee E, Morganstein D, Lipton R. Lost productive time and cost due to common pain conditions in the US workforce. JAMA. 2003;290(18):2443–2454.

2. Directorate General of Health Services (DGHS). Chapter 7 Morbidity Profile. In, Health Bulletin 2014. Management Information System, Directorate General of Health Services, Mohakhali, Dhaka 1212;2014:47–57.

3. Institute of Medicine (IoM). Committee on Advancing Pain Research C, Education. Relieving pain in America: a blueprint for transforming prevention, care, education, and research.Washington, DC: National Academies Press. 2011.

4. Lippe PM, Brock C, David J, Crossno R, Gitlow S: The First National Pain Medicine Summit–final summary report. Pain Med. 2010;11:1447–1468,

5. Tauben DJ, Loeser JD. Pain education at the University of Washington School of Medicine. J Pain. 2013;14(5):431–437.

6. Doorenbos AZ, Gordon DB, Tauben D, Palisoc J, Drangsholt M, Lindhorst T, et al. A blueprint of pain curriculum across prelicensure health sciences programs: one NIH Pain Consortium Center of Excellence in Pain Education (CoEPE) experience. J Pain. 2013;14(12):1533–1538.

7. Bond M. A decade of improvement in pain education and clinical practice in developing countries: IASP initiatives. Br J Pain. 2012;6:81–84.

8. Miles S, Kellett J, Leinster SJ. Medical graduates’ preparedness to practice: a comparison of undergraduate medical school training. BMC Med Educ.2017;17:33

9. Pilowsky I. An outline curriculum on pain for medical schools. Pain. 1988;33(1):1–2.

10. Briggs EV, Battelli D, Gordon D, Kopf A, Ribeiro S, Puig MM, et al. Current pain education within undergraduate medical studies across Europe: Advancing the Provision of Pain Education and Learning (APPEAL) study. BMJ Open. 2015;5(8):e006984.

11. Briggs EV, Carr EC, Whittaker MS. Survey of undergraduate pain curricula for healthcare professionals in the United Kingdom. Eur J Pain. 2011;15(8):789–795.

12. Mezei L, Murinson BB. Johns Hopkins Pain Curriculum Development T. Pain education in North American medical schools. J Pain. 2011;12(12):1199–1208.

13. Phillips DM. JCAHO pain management standards are unveiled. Joint Commission on Accreditation of Healthcare Organizations. JAMA. 2000;284(4):428–429.

14. Miró J, Castarlenas E, Solé E. et al. Pain curricula across healthcare professions undergraduate degrees: a cross-sectional study in Catalonia, Spain. BMC Med Educ. 2019; 19:307

15. Epstein RM, Hundert EM. Defining and assessing professional competence. JAMA 2002;287:226–235.

16. Johora F, Rahman MS. Pharmacology education in the perspective of pharmaceutical promotion: Bangladesh experience. Bangabandhu Sheikh Mujib Medical University Journal 2019;12(3):128–132.

17. Watt-Watson J, McGillion M, Hunter J, Choiniere M, Clark AJ, Dewar A, et al. A survey of prelicensure pain curricula in health science faculties in Canadian universities. Pain Res Manag. 2009;14(6):439–44.

18. Leegaard M, Valeberg BT, Haugstad GK. Survey of pain curricula for healthcare professionals in Norway. Vard Nord Utveckl Forsk. 2014;34(1):42–45.

19. Shipton EE, Bate F, Garrick R, Steketee C, Visser EJ. Pain medicine content, teaching and assessment in medical school curricula in Australia and New Zealand. BMC Med Educ. 2018;18:110.

20. Johnson M, Collett B, Castro-Lopes JM. The challenges of pain management in primary care: a pan-European study. J Pain Res. 2013;6:393–401.

21. Leung SF, Mok E, Wong D. The impact of assessment methods on the learning of nursing students. Nurs Educ Today. 2008;28:711–719.

22. Shipton EE, Steketee C, Bate F, Visser EJ. Exploring assessment of medical students’ competencies in pain medicine-A review. Pain Rep. 2018;12;4(1):e704

